# Visual explanations for the detection of diabetic retinopathy from retinal fundus images

**DOI:** 10.1101/2022.07.06.22276633

**Authors:** Valentyn Boreiko, Indu Ilanchezian, Murat Seçkin Ayhan, Sarah Müller, Lisa M. Koch, Hanna Faber, Philipp Berens, Matthias Hein

## Abstract

In medical image classification tasks like the detection of diabetic retinopathy from retinal fundus images, it is highly desirable to get visual explanations for the decisions of black-box deep neural networks (DNNs). However, gradient-based saliency methods often fail to highlight the diseased image regions reliably. On the other hand, adversarially robust models have more interpretable gradients than plain models but suffer typically from a significant drop in accuracy, which is unacceptable for clinical practice. Here, we show that one can get the best of both worlds by ensembling a plain and an adversarially robust model: maintaining high accuracy but having improved visual explanations. Also, our ensemble produces meaningful visual counterfactuals which are complementary to existing saliency-based techniques. Code is available under https://github.com/valentyn1boreiko/Fundus_VCEs.

## 1 Introduction

In many medical domains, deep learning systems have been shown to perform close to or even better than domain experts in detecting disease from images [21]. For clinicians and patients to trust such systems in practice, they need to be interpretable [14,15]. Current techniques for interpreting model decisions, however, have critical shortcomings. For instance, post-hoc interpretability techniques such as saliency maps are often used to generate explanations for a classifier’s decision. These have been evaluated for clinical relevance, e.g. in ophthalmology [2, 5, 33], with some methods producing more meaningful visualizations than others. As DNNs can rely on spurious features and are not necessarily learning all class-relevant features [11,12], saliency maps may also have limited usefulness in clinical settings [2, 28]: for standard classifiers they sometimes just highlight high-frequency components of an image [5]. Especially for healthy cases, these are often hard to interpret during screening for timely intervention.

Interestingly, models trained to provide inherent robustness against adversarial attacks [7, 22], have also been shown to yield better saliency maps [10, 23]. Also, these robust models allow to generate visual counterfactual explanations (VCEs) [3, 6], an alternative image-wise interpretability technique that shows the minimal changes necessary to maximize the confidence of the classifier in a desired class (Fig. 1). But, the gain of these models in adversarial robustness comes at the price of a loss in accuracy [32, 35] which is unacceptable especially in medical applications. Thus, adversarially robust models have not seen widespread use in practice.

**Fig. 1:**
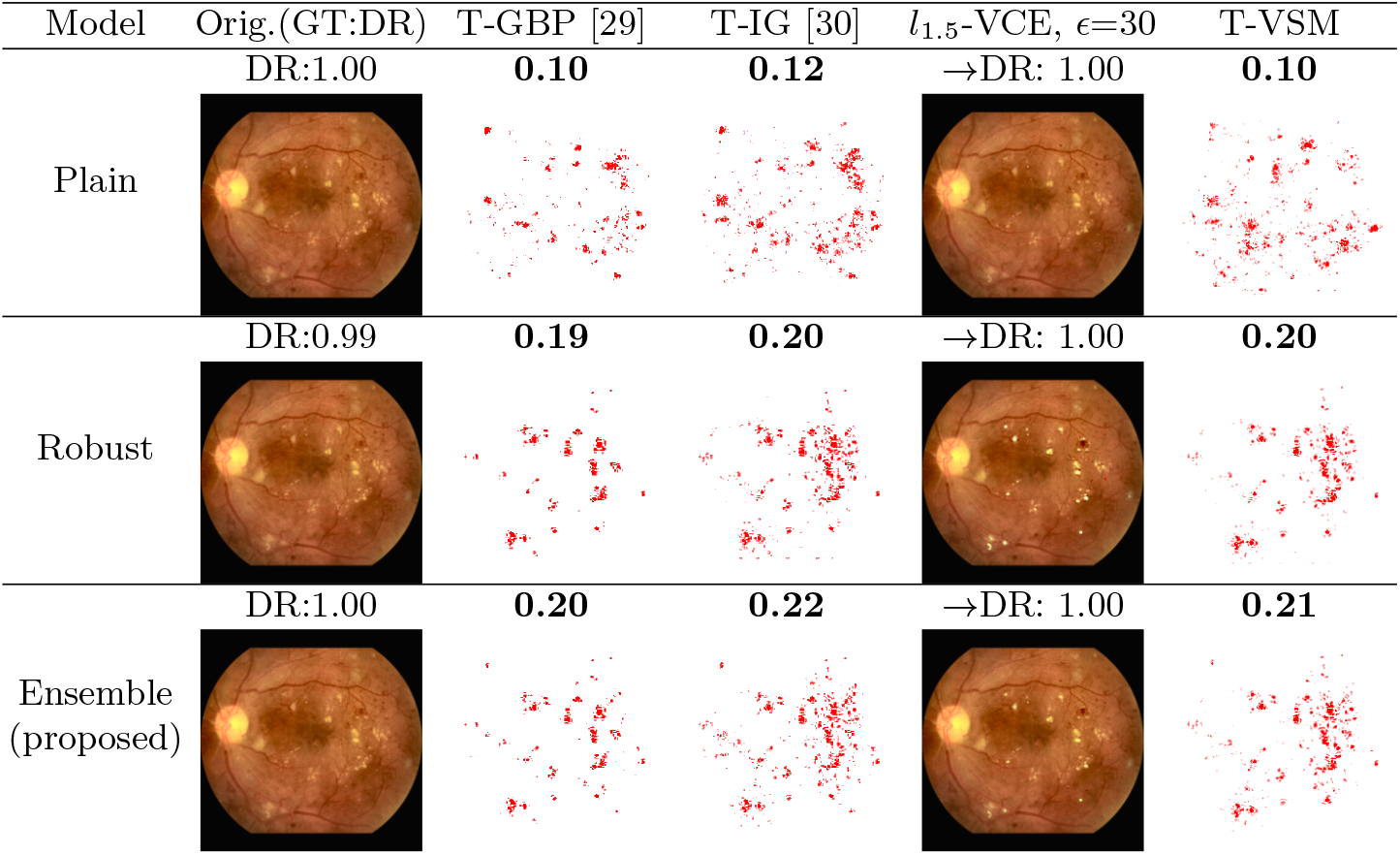
Visual explanations of decisions are better for robust and ensemble models than for plain models, as shown by intersection over union (IoU) between saliency maps (P) and ground truth (GT) masks 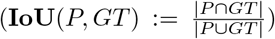 (in bold). We show an image correctly classified as DR (left), post-hoc explanations for the decision using thresholded Guided Backprop (T-GBP), Integrated Gradients (T-IG) and visual counterfactual examples (VCEs) for enhancing the classifiers’ confidence into DR as well as the corresponding saliency map: thresholded VCE Saliency Map (T-VSM). Numerical evaluation of these maps in comparison to the ground truth segmentation can be found in Tab. 2.

Here we show that an ensemble of a plain and an adversarially robust model yields improved saliency maps and allows for the computation of VCEs to further explore the basis of the model’s decision. Further, it achieves almost the same accuracy as the plain model. We demonstrate this new approach to explainability for medical image classifiers for the case of diabetic retinopathy (DR) detection from retinal fundus images and propose a new type of the saliency map.

## 2 Methods

### 2.1 Datasets

We used three publicly available datasets of retinal fundus images for which DR grades were available: the Kaggle DR detection challenge data [1] for method development and main results, the Messidor dataset [9] for additional external validation, and a portion of the Indian Diabetic Retinopathy Image Dataset (IDRiD) [26] for quantitative evaluation of visual explanations, as these data additionally had DR lesion annotations at pixel level. We pre-processed the images using contrast limited adaptive histogram equalization (CLAHE) [36], and by tightly cropping the circular mask of the retinal fundus, which was detected by iterative least-squares fitting of a circular shape to image edges. For the Kaggle dataset, we filtered out poor quality images using an ensemble of EfficientNets [31] trained on the ISBI2020 challenge dataset^1^. This quality filtering model achieved 87.50% accuracy for image gradability. After quality filtering, the resulting dataset contained 45, 923 images (at a final resolution of 224 × 224 pixels): 33, 783 in class ‘no DR’, 3, 598 in ‘mild DR’, 6, 765 in ‘moderate DR’, 1, 186 in ‘severe DR’ and 591 in ‘proliferative DR’. The Messidor dataset contained 1200 retinal fundus images, and the IDRiD 81 images along with annotations for microaneuryms, haemorrhages, hard and soft exudates. We combined the annotations of these lesion types to obtain a single ground truth mask.

### 2.2 Plain, robust and ensemble models

As mild DR is a transitional stage between no DR and moderate-to-advanced stages of DR [34], these images lead to high uncertainty in decisions of both DNNs and clinicians [4]. Therefore, to obtain a clear separation of ‘no DR’ and DR classes, we excluded the ‘mild DR’ cases. We then trained binary classifiers *f* : ℝ^*d*^ → ℝ^2^ to predict whether a fundus image *x* was in the ‘no DR’ class or belonged to moderate-to-advanced stages of DR, with 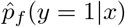 indicating the predicted probability of disease. We used 75% of the Kaggle data for training, 15% for validation, 4% for temperature scaling [16] and 6% for testing.

For the plain model we used a ResNet-50 [17] which was trained with cross-entropy loss. We used batch size of 128, with oversampling of the DR cases to account for class imbalance. We first trained the model for 500 epochs with learning rate of 0.01 and a cosine learning rate schedule. This model was further fine-tuned for 3 epochs with a cyclic triangle schedule for one cycle. We chose the model with the best balanced accuracy on the validation set.

The robust model used the same architecture but was trained using TRADES [35] for *l*_2_-adversarial robustness, where one minimizes for the given training set 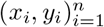 the objective:

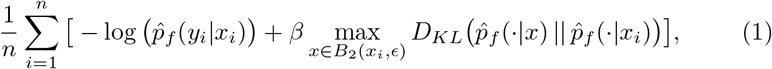

where *D*_*KL*_ denotes the Kullback-Leibler divergence, 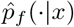 is the predicted probability distribution over the classes at *x, β* controls the trade-off between adversarial and plain training schemes, and 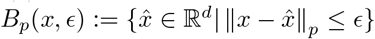. For training we used *p* = 2 and *ϵ* = 0.25 and set *β* = 6.

In our experience, tuning *β* down during training can increase accuracy but negatively affects interpretability. Hence, we built the following ensemble of plain and robust models, which preserves both accuracy and interpretable gradients for the given *β*:

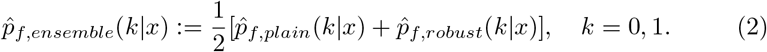

As saliency methods often require logits *f* instead of probabilities, we defined logits for the ensemble as 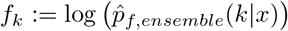. All models are calibrated via temperature scaling by minimizing the expected calibration error [16].

Experiments were done on an Nvidia Tesla V100 GPU with 32GB RAM, using PyTorch. Code for pre-processing and training as well as the trained models will be available upon acceptance.

### 2.3 Generating visual counterfactual explanations (VCEs)

Following [6], a VCE 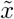 should have high probability 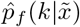 in a chosen class *k* (”validity”). It should be similar to the starting image *x*_0_ (”sparsity”) and close to the data manifold (”realism”). For generating an *l*_*p*_-VCE 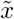 for a classifier *f* we solved

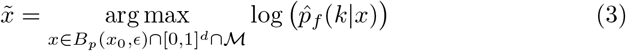

where *ℳ* is the mask for the region of the eye obtained by our pre-processing. The formulation of VCEs suggests that some “robustness” is required as Eq. 3 is similar to the formulation of adversarial examples [6]. Compared to saliency maps the advantage of VCE is that the generated images are purely based on the behavior of the classifier. We used adaptive projected gradient descent (APGD) [7] and Frank-Wolfe [19, 24] based schemes as optimizers. APGD requires projections onto *l*_*p*_-balls which are available in closed form for *l*_2_ and *l*_∞_ or can be computed efficiently for *l*_1_ [8]. However, for *p* ∉ {1, 2, ∞}, there is no such projection available and thus we used for the generation of *l*_*p*_-VCEs the Auto-Frank-Wolfe scheme of [6].

### 2.4 Saliency maps

We used Guided Backprop (GBP) [29] and Integrated Gradients (IG) [30] from a public repository [25] to generate saliency maps for the models’ decisions. GBP and IG are among the best saliency techniques for DR detection [5, 33]. Based on our VCEs, we also introduced the VCE Saliency Map (VSM) as the difference between VCE and the original image. For all saliency methods, we used absolute saliency values summed over color channels in order to better cover salient regions [5]. Then, saliency scores were normalized to [0, 1] via minmax normalization and thresholded at the *τ* -quantile for sparsity. The threshold *τ* was optimized for each method on 40 out of 81 images in the IDRiD dataset by computing the intersection over union (IoU) with respect to the pixel-wise annotation of DR lesions. This yielded *τ* = 0.98 for GBP, *τ* = 0.96 for both IG and VSM. For the VSMs we additionally optimized over the norm *p ∈* { 1.5, 2, 4 } and different *ϵ* per norm and found *p* = 1.5, *ϵ* = 30 to be the best.

### 2.5 Model evaluation

We evaluated the performance of models on the Kaggle test set and Messidor images using accuracy (acc.), and balanced accuracy (bal. acc., mean of TPR and TNR). Additionally, we reported *l*_2_-robust accuracy (rob. acc.) for a perturbation budget of *ϵ* = 0.1 which we evaluated using 9 restarts of 100 iterations of APGD [7] maximizing the confidence in the wrong class. The robust accuracy is the fraction of test inputs where the decision could not be changed by the attack.

For a quantitative evaluation of our visual explanations, we used the 41 images on which *τ* had not been optimized from the IDRiD dataset. Tab. 2 shows the mean IoU for all models and saliency techniques (including T-VSMs for different *p*-norms) with the pixel-level DR lesion annotations.

This evaluation indicates that the saliency maps derived from VCEs are on par with state-of-the-art techniques, such as GBP and IG. However, VCEs go beyond those techniques as they can be used to generate images and even animations that illustrate how an image would have to change to affect the prediction of the classifier.

## 3 Results

First, we analyzed the properties of the plain and robust classifiers, and the ensemble introduced in Eq. 2. Then, we explored VCEs as an alternative for explaining classifier decisions and studied the sparsity-realism trade-off for VCEs. Finally, we show the effect of different perturbation budgets on VCEs.

### 3.1 Ensembling plain and adversarially trained DNNs

We found that the plain model achieved good standard and balanced accuracy for classifying DR from fundus images (Tab. 1), but with comparably low robust accuracy (see Sec. 2.5). In contrast, the robust classifier achieved high robust accuracy, but suffered a large drop in accuracy of more than 10-20%. Interestingly, and in line with the literature [10, 23], the saliency maps of the robust model were much better than those of the plain model (Tab. 2, Fig. 1) for both of the tested saliency methods, Guided Backprop (GBP) and Integrated Gradients (IG). In fact, the saliency maps of the plain classifier were of rather low quality, focusing on less prominent disease-related regions of the image (Fig. 1).

**Table 1:** Evaluation of plain and robust classifier and their ensemble in terms of standard, balanced and *l*_2_-robust accuracy. The ensemble maintains the accuracy but gains sufficient robustness required for better interpretability (see Tab. 2).

|  | Kaggle |  |  | Messidor |  |  |
| --- | --- | --- | --- | --- | --- | --- |
|  | acc. | bal. acc. | rob. acc. | acc. | bal. acc. | rob. acc. |
| Plain | 89.5 | 85.8 | 15.2 | 89.5 | 89.5 | 20.6 |
| Robust | 78.4 | 71.6 | 66.6 | 66.1 | 66.5 | 60.9 |
| Ensemble | 89.7 | 85.2 | 19.4 | 87.9 | 87.9 | 24.4 |

**Table 2:**
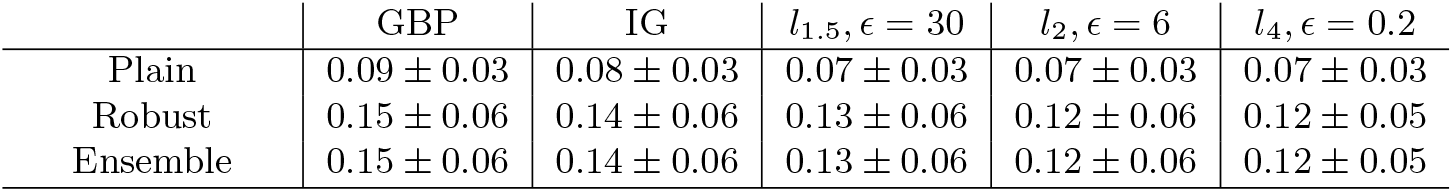
Evaluation of saliency maps and T-VSMs on IDRiD. The IoU-score of the ensemble is higher than for the plain model for all interpretability methods including VCEs (higher is better, mean ± std).

| | GBP | IG | $l_{1.5}, \epsilon = 30$ | $l_2, \epsilon = 6$ | $l_4, \epsilon = 0.2$ |
| --- | --- | --- | --- | --- | --- |
| Plain | $0.09 \pm 0.03$ | $0.08 \pm 0.03$ | $0.07 \pm 0.03$ | $0.07 \pm 0.03$ | $0.07 \pm 0.03$ |
| Robust | $0.15 \pm 0.06$ | $0.14 \pm 0.06$ | $0.13 \pm 0.06$ | $0.12 \pm 0.06$ | $0.12 \pm 0.05$ |
| Ensemble | $0.15 \pm 0.06$ | $0.14 \pm 0.06$ | $0.13 \pm 0.06$ | $0.12 \pm 0.06$ | $0.12 \pm 0.05$ |

We found that an ensemble of the plain and robust models (Eq. 2) combined their advantages: It had about equal standard and improved robust accuracy compared to the plain model (Tab. 1) and its saliency maps were as good as those of the robust model (Tab. 2, Fig. 1).

### 3.2 VCEs as an alternative to saliency maps

We next explored VCEs (Eq. 3) as an alternative for explaining classifier decisions. The properties of the VCEs depend on the chosen model for the perturbation, which in this paper was always an *l*_*p*_-ball, and the perturbation budget in form of the radius of *l*_*p*_-ball. Small values of *p* close to one lead to sparse changes whereas for larger *p* one can realize much more outspread changes affecting larger parts of the image. As discussed in Sec. 2.4 we chose *l*_1.5_-VCEs of radius *ϵ* = 30 as they produced the best quality of T-VSMs. We found that the robust model and the ensemble allowed for the computation of realistic VCE (Eq. 3, Fig. 1). T-VSMs (see Sec. 2.4) also provided good explanations for the classifiers’ decision (Tab. 2), highlighting exudates and haemorrhages. In contrast, the VCE of the plain model was not very meaningful as its main changes were only vaguely related to the diseased regions.

### 3.3 Sparsity versus Realism of VCEs

We then analyzed the effect of different perturbation models in terms of different *l*_*p*_-balls (Fig. 2). We first studied the VCEs for enhancing the correct decision for a DR image. We found that the changes of *l*_1.5_-perturbation model were sparser and thus looked more cartoon-like than for *l*_4_. The VCEs of the *l*_4_ model appeared much more natural although they even introduced new diseased regions not present in the original image. Thus the classifier seems to have picked up certain disease signs very well and can integrate even new disease patterns in a natural fashion into fundus images. We next studied the VCE for changing the decision of the classifier to ‘no DR’. Here, all *l*_*p*_-perturbation models attempted to “smooth out” the main lesions as well as the exudates. This provides complementary evidence that the classifier picked up the right disease signal in the data. Note that the artefact around the optic nerve was not changed in the VCE, showing that the classifier has correctly identified it as a feature which is not discriminatory for the disease decision. Not all VCEs, however, provided by our method are perfectly realistic: for example, the algorithm often tried to cover lesions with vessels when creating a VCE turning a diseased image into a healthy one. Further failure cases are discussed in App. A.

**Fig. 2:**
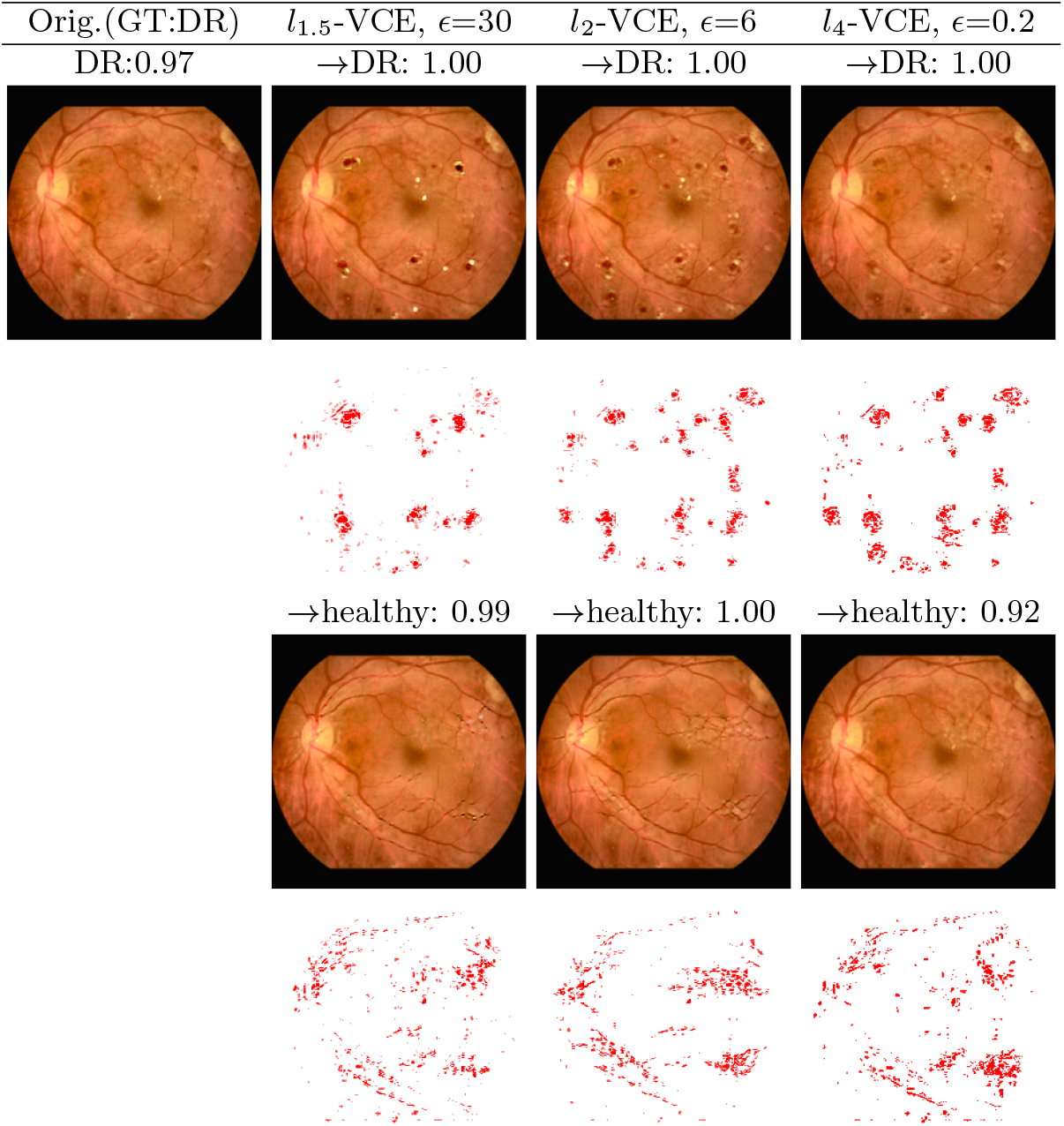
VCEs for the ensemble with varying degree of sparsity: *p* ∈ { 1.5, 2, 4 }. For a correctly classified DR image, we show VCEs when transformed further into the DR or the healthy class. Below VCEs, T-VSMs are shown. The VCE radius was adapted to the sparsity condition. In addition, the confidence of the classifier is reported above the image.

### 3.4 VCEs for different budgets

Finally, we investigated how the VCEs changed with increasing budget parameterized with *ϵ* (Fig. 3). We found that an increasing number of new lesions were introduced for both the sparse *l*_1.5_-VCE as well as the realistic *l*_4_-VCE, when increasing the budget for more DR evidence. Here, the difference between the two models — that *l*_4_-VCEs appeared more realistic — became even more clear. When generating VCEs for turning the diseased image into an healthy one, also increasingly large regions of lesions were covered, e.g. through artificial vessels. Such VCE with different budgets could be useful to generate gradual changes in either directions, providing good intuitions for a classifiers decision.

**Fig. 3:**
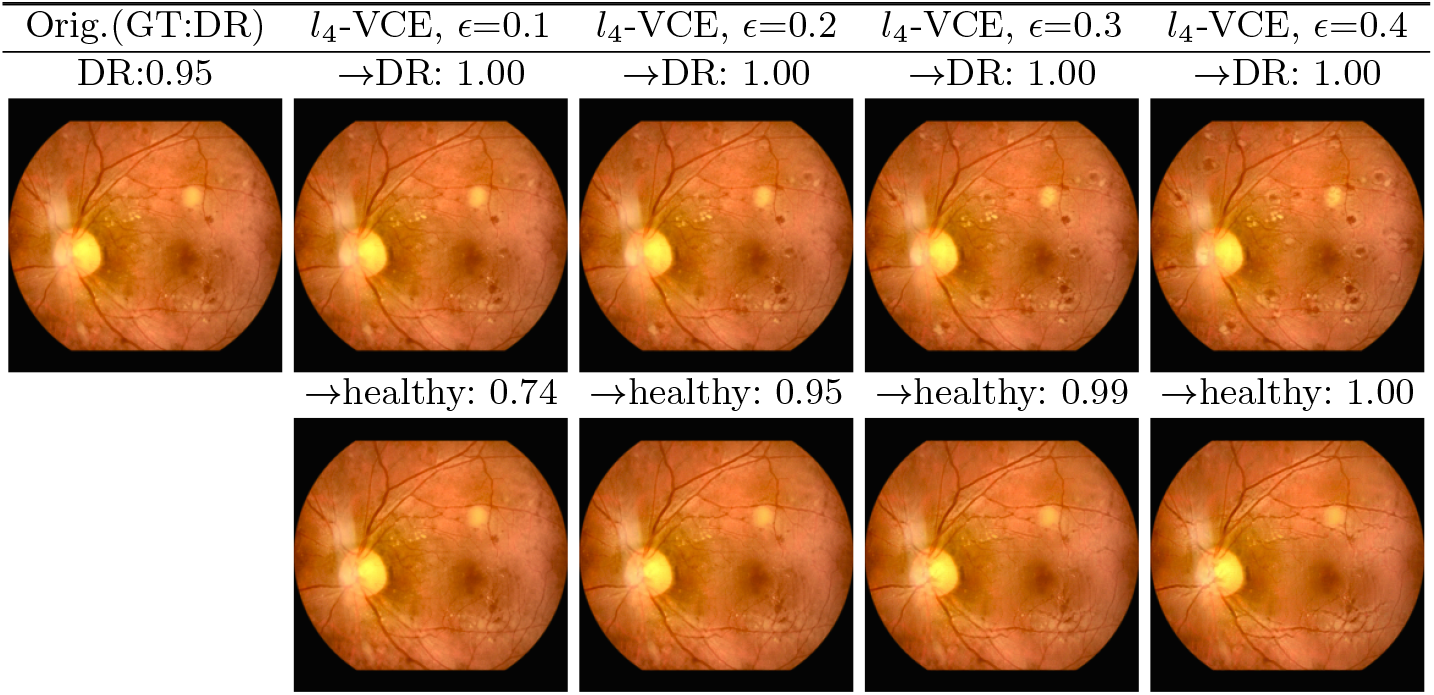
VCEs show increasingly strong modification for different radii. For one correctly classified DR image, we show for the ensemble the *l*_4_-VCEs for *ϵ* ∈ {0.1, 0.2, 0.3, 0.4} when transforming into the DR and healthy class, respectively.

## 4 Discussion

We showed that the ensemble of plain and robust models can preserve accuracy of plain models, yet provide better visual explanations. In agreement with the literature [10, 23], the resulting saliency maps highlight clinically relevant lesions more reliably. Therefore, the explanations obtained for diseased images are often satisfying, while those for healthy images are less so — showing the absence of lesions is difficult in this framework. The ensemble model allowed us to compute also realistic VCEs [6], to yield interpretable explanations of the classifier’s decision, pinpointing the features in the image the classifier picks up on.

In related work, iterative augmentation of saliency maps has been used to improve saliency-based visual explanations [13]. Also, VCEs have been generated using GANs [20] (no models/code is available) but the advantage of our VCE is that they depend only on the classifier and thus there is no danger that the prior of the GAN “hides” undesired behavior of the classifier. Finally, models interpretable-by-design such as BagNets [18] have been advocated for medical imaging tasks [27]. As many high-performing DNNs do not fall into this category, we view our work as complementary.

We believe realistic VCEs and derived T-VSMs will be a useful tool to better understand the behavior of DNN-based classifiers in medical imaging, in particular when gradually morphing an image from one class to the other which is the main complementary strength of VCEs compared to saliency maps. As the sparseness and the degree of changes allowed can be precisely controlled, it is straightforward to yield more or less natural VCEs. Even extreme and therefore less natural VCEs can be useful, as they provide a “cartoon” version of what the classifier believes the disease looks like.

## Data Availability

All data produced are available online at

https://www.kaggle.com/c/diabetic-retinopathy-detection

https://www.adcis.net/en/third-party/messidor/

https://ieee-dataport.org/open-access/indian-diabetic-retinopathy-image-dataset-idrid

https://isbi.deepdr.org/challenge2.html

## Acknowledgement

We acknowledge support by the German Ministry of Science and Education (BMBF, 01GQ1601 and 01IS18039A) and the German Science Foundation (BE5601/8-1 and EXC 2064, project number 390727645). The authors thank the International Max Planck Research School for Intelligent Systems (IMPRS-IS) for supporting I.I.

## A Failure cases

**Fig. 4:**
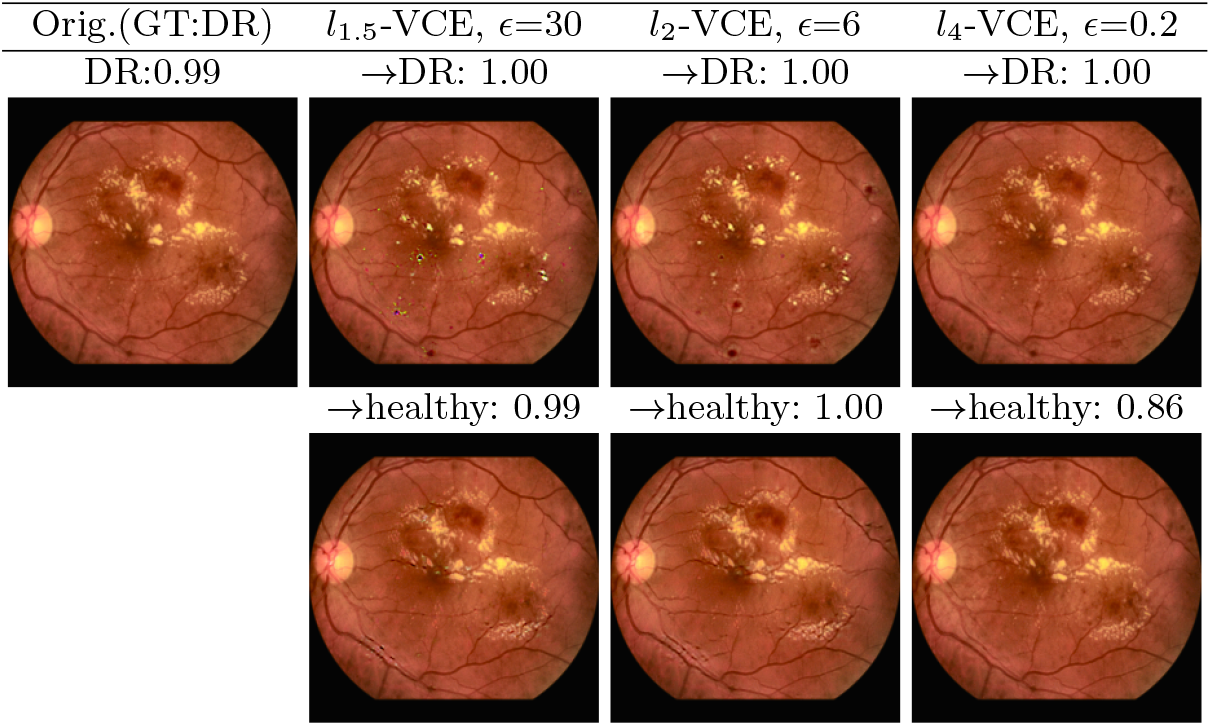
Failure: when transforming to DR using ensemble, *l*_1.5_-VCE has visible artifacts, unlike *l*_2_-,*l*_4_-VCEs.

**Fig. 5:**
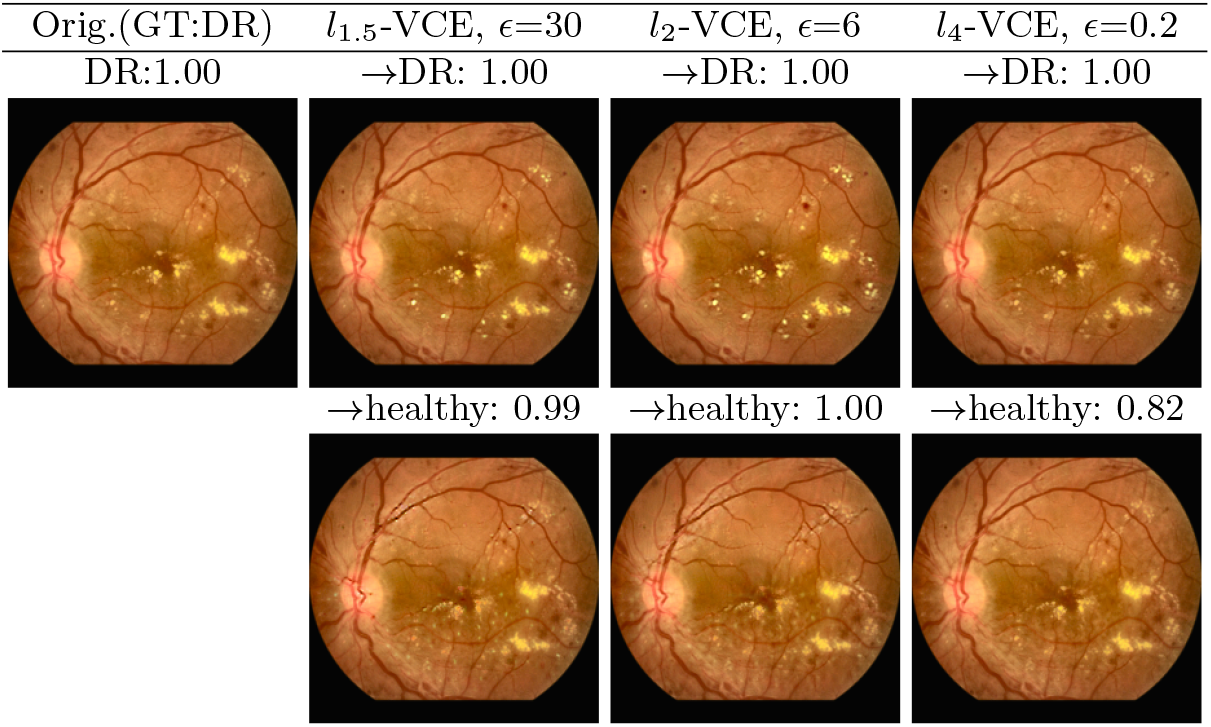
Failure: when transforming to healthy using ensemble, *l*_2_- and *l*_1.5_-VCEs have visible artifacts (yellow spots), unlike *l*_4_-VCE.

## B Further examples of VCEs

**Fig. 6:**
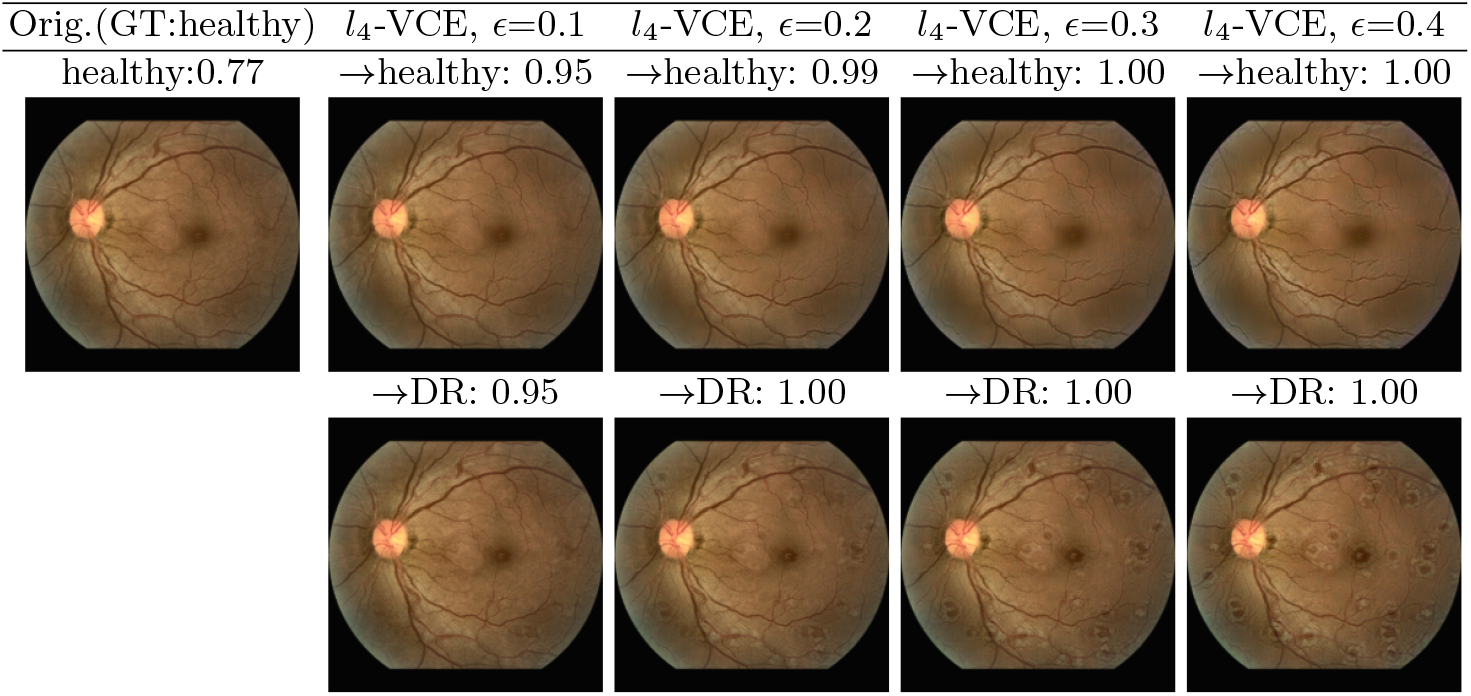
For one correctly classified healthy image, we show for the ensemble the *l*_4_-VCEs for *ϵ∈* { 0.1, 0.2, 0.3, 0.4 } when transforming into the healthy and DR class, respectively.

**Fig. 7:**
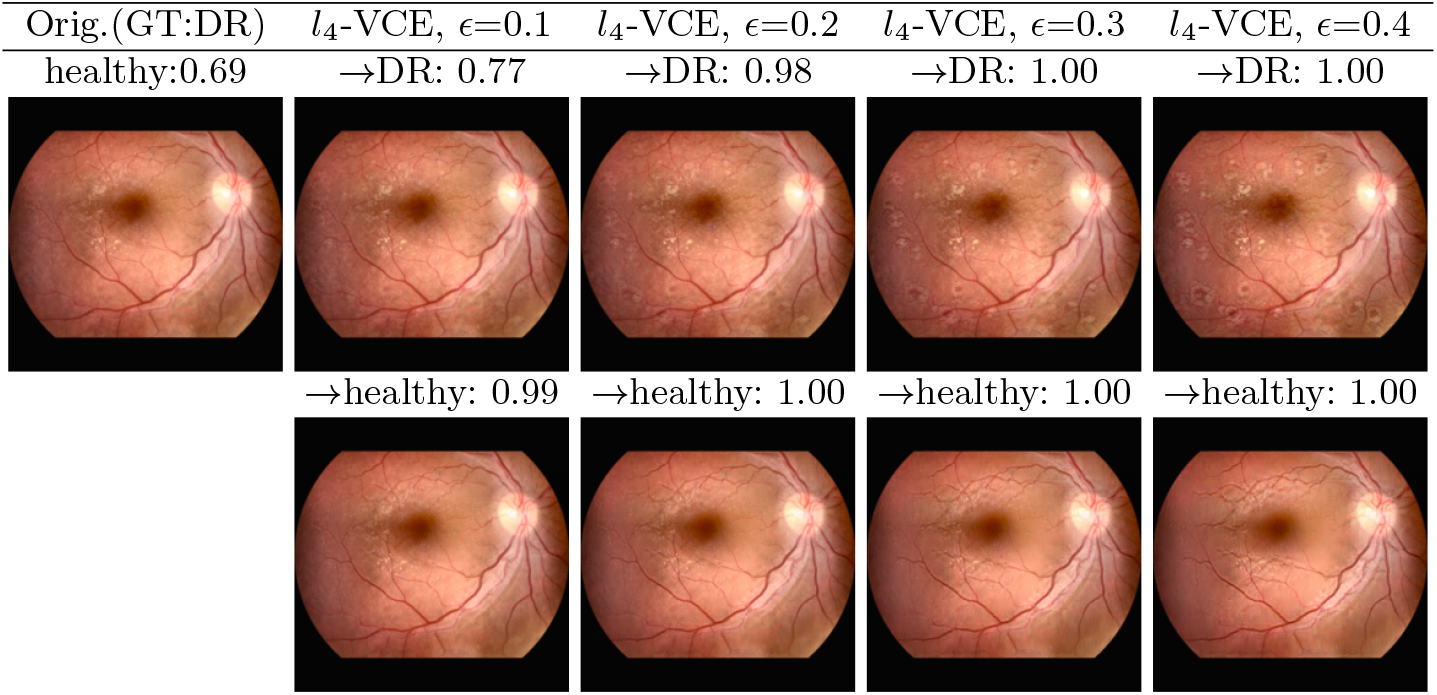
For one wrongly classified DR image, we show for the ensemble the *l*_4_- VCEs for *ϵ* ∈ { 0.1, 0.2, 0.3, 0.4 } when transforming into the DR and healthy class, respectively.

## Footnotes

1 https://isbi.deepdr.org/challenge2.html

